# Prevalence of ocular manifestations in cutaneous rosacea: Protocol for a systematic review and meta-analysis

**DOI:** 10.1101/2024.01.31.24301198

**Authors:** Kristina Nazzicone, Ryan H. Kirkpatrick, Aleksandar Biorac, Anneke R. Froentjes, Sonja Molin, Sarah Simpson

**Affiliations:** Department of Medicine, Queen’s University, Kingston, Ontario, Canada; Centre for Neuroscience Studies, Queen’s University, Kingston, Ontario, Canada; Division of Dermatology, Queen’s University, Kingston, Ontario, Canada; Department of Ophthalmology, Queen’s University, Kingston, Ontario, Canada

**Keywords:** ocular rosacea, rosacea, meta-analysis, systematic review, protocol

## Abstract

Introduction: Rosacea is a chronic inflammatory skin condition with concomitant ocular manifestations and neurogenic symptoms. Ocular rosacea poses a particular diagnostic challenge as its signs and symptoms often overlap with other ocular pathologies. Cutaneous and ocular rosacea present together in approximately 21-50% of patients, yet a solid understanding of comorbid ocular and cutaneous symptoms is lacking. Therefore, the present paper outlines a protocol for a systematic review and meta-analysis to describe and quantify the prevalence of ocular rosacea in cutaneous rosacea and each of its subtypes. Methods: This study will follow Preferred Reporting Items for Systematic Reviews and Meta-Analyses (PRISMA) guidelines and be conducted using the systematic review software, Covidence. To determine inclusion, screening will occur at two levels (title and abstract-only followed by full-text) and will be completed separately by two authors. Primary research studies on ocular rosacea in adult cutaneous rosacea published in all languages and geographic regions until November 2023 will be reviewed for inclusion. Data pertaining to overall prevalence of ocular rosacea across and within cutaneous rosacea subtypes, mean age, sex, gender, ethnicity, socioeconomic status, time to diagnosis, time to treatment, and prevalence of comorbid conditions will be extracted. For each included study, the Grading of Recommendations, Assessment, Development, and Evaluations (GRADE) framework will be applied to assess study quality. Conclusion: To the authors’ knowledge, this will be the first systematic review and meta-analysis comparing the prevalence of ocular rosacea in the various cutaneous subtypes in an adult population. By addressing this knowledge gap, this study aims to provide clear and easily interpretable data to aid in the early diagnosis and treatment of ocular rosacea. This study is registered in the International Prospective Register of Systematic Reviews (PROSPERO ID# CRD42023475026).

**Key Message:** Protocol for a systematic review and meta-analysis investigating the prevalence of ocular manifestations in adult rosacea

## INTRODUCTION

Rosacea is a chronic inflammatory skin condition with a global prevalence of approximately 5.46% of the general population and 2.39% of dermatologic outpatients (1). The prevalence of rosacea is almost 50% higher in females (5.41%) compared to males (3.90%;1). It affects the face (cheeks, forehead, eyes, nose, and chin) and has a typical onset between 30 and 50 years of age (2).

The psychosocial impact of rosacea should not be underestimated. In a survey conducted by the National Rosacea Society, 75% of respondents reported lowered self-esteem directly related to their rosacea with 52% of respondents explicitly reporting that they had avoided face-to-face interactions due to their rosacea (3). The impact of rosacea on quality of life has been compared to that of leg ulcers and vitiligo (4). Through the retrospective study of American insurance records, individuals with rosacea were found to spend $735 USD more on health-related costs per year than matched controls, with $276 USD per year being specifically related to their rosacea (5).

The diagnosis of rosacea is made clinically. Symptoms may be subtle or overlap with other common conditions, which is often the case in ocular manifestations of rosacea (6). Rosacea often presents with facial flushing, erythema (persistent or episodic), papules and/or pustules, telangiectasia, phymata, ocular manifestations, and neurogenic symptoms (2,7,8). Historically, rosacea has been underdiagnosed and underreported in persons with darker skin tones (Fitzpatrick skin types IV, V, or VI), presumably due to the lack of well-defined characteristics of the disease, and the hallmark sign of erythema being more challenging to appreciate (9).

The etiology and pathophysiology of rosacea is not completely understood, yet thought to be caused by a combination of genetic factors, neurovascular and immune dysregulation, and extrinsic triggers; namely microorganisms and environmental exposures (8). Ultraviolet (UV) exposure is known to trigger rosacea episodes, though it is not clear if UV exposure is associated with its initial development or etiology (10). *Demodex* mites are seen in larger quantities in rosacea-affected skin, yet it is unclear if the higher numbers of these microorganisms exist as a consequence of rosacea or are a contributory cause (11–13). More recent research has highlighted the association between rosacea and other systemic diseases including cardiovascular (14–16), gastrointestinal (17–19), neurologic (20,21) and other autoimmune conditions (22). A possible genetic influence has also been suggested as rosacea has a higher incidence in those with a family history of the disease (23). This observation is further supported by the identification of specific human leukocyte antigen (HLA) loci in patients with rosacea (23).

Rosacea can be separated into four clinical subtypes: erythematotelangiectatic, papulopustular, phymatous, and ocular. The reported proportion of individuals with comorbid cutaneous and ocular manifestations of rosacea varies in the literature and ranges from 21-50% (24,25). It has also been suggested that ocular manifestations of rosacea may appear prior to cutaneous features of the disease (25). Similar to cutaneous rosacea, ocular rosacea is a clinical diagnosis based on signs and symptoms. Common eye manifestations include: dryness, blurred vision, foreign body sensation, burning/stinging, photophobia, blepharitis, meibomian gland dysfunction, and eyelid chalazion (6,26). In serious cases, rosacea can lead to vision loss through corneal inflammation, sclerokeratitis, and cicatricial conjunctivitis (6,27).

Primary research on rosacea often reports on cutaneous and ophthalmic manifestations, however, to the authors’ knowledge, there have been no reviews consolidating the literature through meta-analysis. There have, however, been narrative reviews and commentaries (26–28) published on ocular rosacea in the past few years, lending support to its clinical interest and importance. Therefore, identifying the prevalence of ocular rosacea in those with cutaneous rosacea is of clinical relevance.

Ocular rosacea is a disease that exists at the intersection of two medical specialties: dermatology and ophthalmology. Thus, clear knowledge of the likelihood of ocular involvement in cutaneous rosacea and within each cutaneous rosacea subtype is important for prompt and appropriate referral to ophthalmology, or the reverse, if the ocular manifestations present first without cutaneous disease. Knowledge of the relationship between ocular and cutaneous rosacea can help physicians better assess and predict the natural history of rosacea and provide a framework for which patients are more likely to present with both cutaneous and ocular symptoms. This has the potential to improve patient care by reducing the time from symptom onset to diagnosis and, ultimately, treatment.

The present paper outlines the protocol for a systematic review and meta-analysis to quantify the prevalence of ocular rosacea within cutaneous rosacea. The proposed study will review the current body of evidence on ocular manifestations in adult cutaneous rosacea to answer the following question: what is the prevalence of ocular rosacea in adults diagnosed with a cutaneous rosacea. The primary outcomes are to determine the proportion of individuals with cutaneous rosacea that also have ocular rosacea and to determine whether ocular rosacea is more strongly associated with any of the rosacea subtypes (i.e., is there a higher prevalence of ocular rosacea within erythematotelangiectatic, papulopustular, or phymatous rosacea or is the prevalence of ocular rosacea the same across all rosacea subtypes?). Additional outcomes of interest include the time from symptom onset to rosacea subtype diagnosis, and the time from subtype diagnosis to treatment. To the authors’ knowledge, this will be the first systematic review and meta-analysis comparing the prevalence of ocular rosacea in the various cutaneous subtypes in an adult population. By addressing this knowledge gap, this study aims to provide clear and easily-interpretable data to aid in the early diagnosis and treatment of ocular rosacea.

## METHODS

The present protocol follows the Preferred Reporting Items for Systematic Reviews and Meta-Analysis (PRISMA) guidelines (29,30) and is registered in the International Prospective Register of Systematic Reviews (PROSPERO ID# CRD42023475026).

### ELIGIBILITY CRITERIA

#### Study Types

All primary research studies on rosacea and ocular rosacea will be considered eligible for inclusion in the review. This includes randomized controlled trials, case-control studies, cohort studies, and registered clinical trials. In line with recommendations by Cochrane Systematic Reviews, grey literature including dissertations, theses and conference abstracts will also be assessed for inclusion. Case reports and case series will not be included. To be eligible for inclusion, studies must report on the proportion of adults with rosacea that have ocular rosacea or an ocular involvement suggesting ocular rosacea (i.e., blepharitis, meibomian gland dysfunction).

#### Participant Types

Only studies including adults (aged 18 and above) will be included in the analysis. To be eligible for inclusion, study participants must have a diagnosis of rosacea confirmed by a clinician (i.e., diagnosis cannot be made on self-report symptoms). The receipt of treatment or follow-up care for rosacea will be considered to indicate a clinician diagnosed rosacea. Each included study must report on the proportion of patients with a cutaneous rosacea diagnosis that have either a concurrent diagnosis of ocular rosacea or ocular involvement with signs/symptoms consistent with ocular rosacea. This review will not exclude studies that do not explicitly diagnose patients with “ocular rosacea” because ocular rosacea is sometimes labeled as ocular involvement in rosacea literature, given its nonspecific diagnostic criterion. For example, if a study reports on the proportion of individuals with a cutaneous rosacea that also experience blepharitis, it will be considered to assess an ocular manifestation of rosacea and therefore included within the review. By including studies referring to “ocular rosacea” through signs and symptoms rather than only those that state ocular rosacea, this study aims to encompass literature studying ocular rosacea more comprehensively. Studies will not be excluded based on participant disease severity, comorbidities (ocular, dermatologic, or other), length of disease, or treatment history.

#### Defining Ocular Manifestations/Involvement

All relevant ocular manifestations will be eligible for inclusion. Specific search terms will broadly cover ocular disorders and disturbances along with specific signs and symptoms that are characteristic in ocular rosacea (e.g., photophobia, dry eye, eye burning, meibomian gland disease, meibomian gland dysfunction, lacrimal gland/apparatus disease, blepharitis). In instances where it is unclear whether the ocular symptoms reported may represent ocular rosacea, one of the medical specialists involved within the study (authors SS and SM) will be consulted for clinical expertise.

#### Outcome Measures

To measure the prevalence of ocular rosacea in cutaneous rosacea, various metrics will be extracted from each study (where available). The following data will be extracted from each study deemed eligible for inclusion: overall prevalence of ocular rosacea collapsed across all cutaneous rosacea subtypes, prevalence of ocular rosacea within each cutaneous rosacea subtype separately, mean age, sex, gender, ethnicity, socioeconomic status, and prevalence of comorbid conditions. The following disease characteristics will also be extracted: disease severity, length of time from symptom onset to diagnosis or diagnosis to treatment. Finally, study specific characteristics will also be extracted including publication year, sample size, and country of data collection.

#### Information Sources

Journal articles in all languages and geographic regions published up until November 2023 will be reviewed for inclusion, however searches will only occur in English. The following databases and registries will be used to identify studies: Embase, MEDLINE, PubMed, Web of Science, ProQuest, and Cochrane Central Register of Controlled Trials (CENTRAL).

#### Search Strategy

Literature searches will be conducted such that articles must include one term related to rosacea and another to ocular rosacea or ocular involvement (shown in Table 1). A health sciences librarian was consulted for the development of the search strategy to ensure it appropriately reflected the research question.

**Table 1.** Terms used for literature searches. Screened articles must have one item from “Term A” and one item from “Term B” to be included in title-level screening.

| Term A | Term B |
| --- | --- |
| - Rosacea | - Ocular rosacea<br>- Ocular involvement<br>- Ocular or eye disorder*<br>- Ocular or eye disturbance*<br>- Ocular or eye abnormalit*<br>- Ocular or eye disease*<br>- Ocular or eye dysfunction<br>- Ocular or eye burning<br>- Ocular or eye discomfort<br>- Ocular or eye inflammation<br>- Ocular or eye irritation<br>- Ocular or eye redness<br>- Oculopathy<br>- Ophthalmopathology<br>- Ophthalmopathy<br>- Blepharitis<br>- Meibomian gland disease<br>- Meibomian gland dysfunction<br>- Lacrimal gland disease<br>- Lacrimal apparatus disease<br>- Dry eye<br>- Eye burning<br>- Eye discomfort<br>- Eye inflammation<br>- Eye irritation<br>- Eye redness<br>- Eyelid disease<br>- Ocular surface disease<br>- Photophobia |

#### Study Selection

Original literature searches will be conducted by two authors (RHK and KN) for studies published on or before November 2, 2023. Search results will be exported from each search engine and imported in the systematic review software Covidence (31). Duplicate records will be automatically removed by Covidence and manually removed by screeners within the first stage of screening.

Each search result will be reviewed separately by two of four reviewers (authors RHK, KN, AB, AF) at each stage of screening. Papers will progress through two stages of screening: title and abstract, and full text. If an abstract is not included in search engine export and therefore not imported into Covidence, only the title will be used for the first stage of screening (title and abstract stage) to determine advancement to full-text review. Title and abstract screening will follow a standardized and systematic decision-making process used by each screener to minimize unnecessary conflicts (shown in Fig. 1.). For a paper to progress from one stage of screening to the next, two reviewers must agree on its inclusion. In the instance of discrepancies, a third reviewer will decide on inclusion based on the eligibility criteria stated above.

**Fig. 1.**
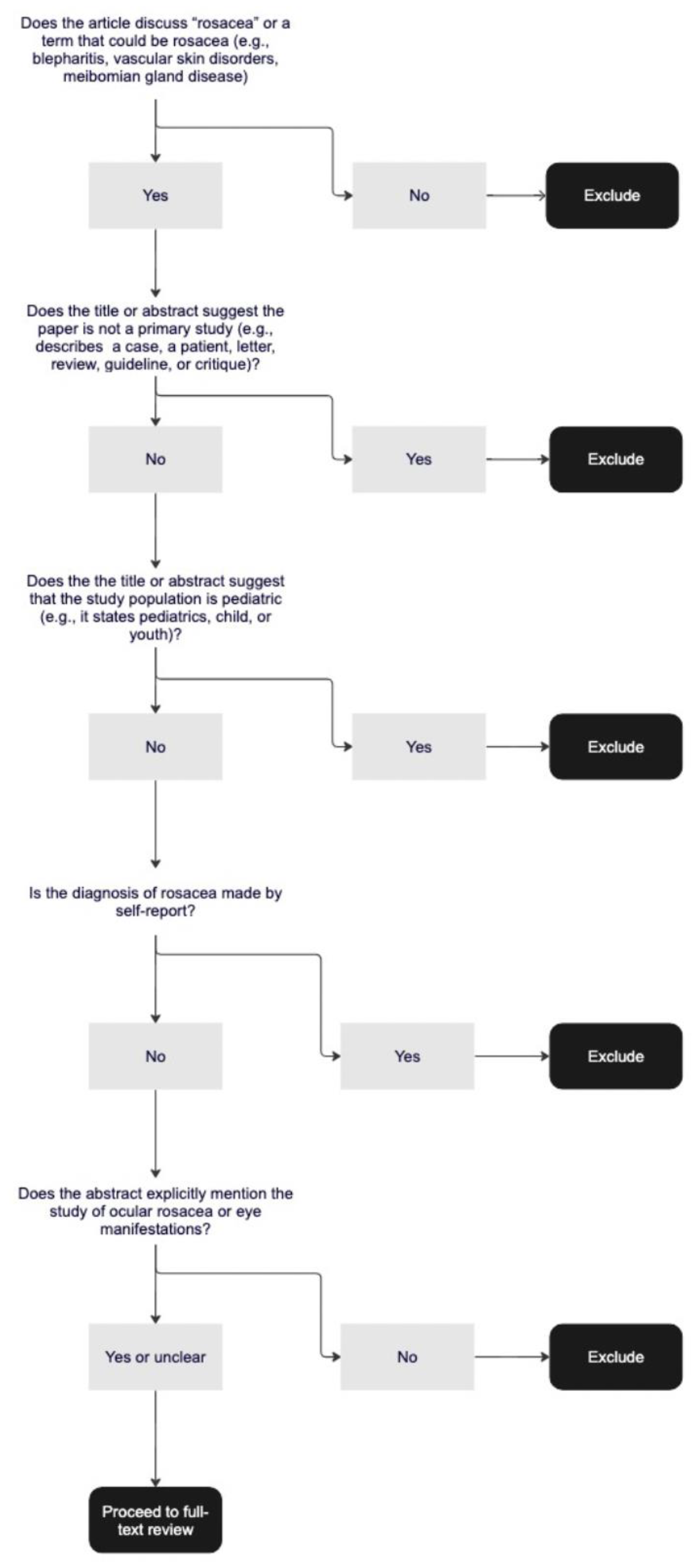
Schema that will be used by reviewers to determine paper advancement from title and abstract-level screening to full-text review.

In the case of multiple publications for a given first or last author, the corresponding author of each paper will be contacted to inquire about overlap of data across papers. If no response is received, only the paper with the highest sample size will be included in the analysis to minimize the likelihood of accounting for the same individual more than once.

#### Data Extraction

Covidence will be used for data extraction. The following items will be extracted from each paper: prevalence of ocular rosacea across rosacea subtypes, prevalence of ocular rosacea within each subtype, age, sex, gender, ethnicity, socioeconomic status, comorbid conditions, disease severity, length of time from symptom presentation to diagnosis, length of time from diagnosis to treatment, publication year, sample size, and country of data collection.

#### Data Transformation

When possible, data will be extracted directly from the papers without any transformation. Should an extracted variable (e.g., age) only be available by group (e.g., age by rosacea subtype), group means will be combined using mean, group sample size, and standard deviation as suggested by the Cochrane Handbook for Systematic Reviews of Interventions (32). If median and interquartile range is reported rather than mean and standard deviation, methods previously published by Wan and colleagues will be used to calculate mean and standard deviation (33).

#### Quality Assessment

The Grading of Recommendations, Assessment, Development and Evaluation (GRADE) framework will be applied to each included study to determine the quality of the evidence used in this review. The primary outcome of each included study will receive a GRADE certainty rating from very low (the true effect is probably markedly different from the estimated effect) to high (the authors have a lot of confidence that the true effect is similar to the estimated effect; 34). Two reviewers will independently determine GRADE certainty ratings and subsequent inter-rater agreement will be calculated. A funnel plot with Peters’ regression will be used to assess publication bias within the grade framework (35)

#### Data Analysis

A PRISMA flow-chart depicting the number of studies included and excluded from the analysis will be developed by Covidence. Should sufficient studies be identified, a meta-analysis will be performed for the prevalence of ocular rosacea in rosacea (i.e., collapsed across subtypes) and each rosacea subtype separately. If insufficient studies are identified for quantitative analysis, findings will be narratively reviewed.

Meta-analyses are anticipated to be performed using the “metaprop” function of the “meta” package (36) within R statistical software (37) however, other software will be considered at the time of analysis. It is anticipated that R will also be used for forest and funnel plot development.

## CONCLUSION

Ocular manifestations of rosacea, or ocular rosacea, has a notable symptom burden and drastic impact on quality of life (38). While individual studies report a prevalence of ocular rosacea in 21-50% of patients with rosacea, it is unclear what its true prevalence within rosacea is, and whether any cutaneous subtypes have a higher prevalence of co-occurring ocular rosacea (25). This lack of understanding is clinically important with regard to the accurate diagnosis and appropriate treatment of ocular manifestations of rosacea. Thus, the present paper outlines a systematic review and meta-analysis to describe the overall prevalence of ocular rosacea in rosacea and within each of the three cutaneous subtypes. Such a review is not only warranted but necessary to consolidate and compare the literature on the subtypes of the disease.

The proposed study acts as the first to address a clinically relevant knowledge gap in the understanding and characterization of rosacea. The proposed study also presents an opportunity to describe and quantify any delay in diagnosis and resultant delay in treatment that might exist in this demographic of patients. By conducting this review and addressing these knowledge gaps, this study aims to contribute high-quality, easily-interpretable data to aid clinicians in better understanding co-morbid ocular and cutaneous rosacea. It is anticipated that findings from this study will be consolidated and described according to PRISMA guidelines, submitted to peer-reviewed journals, and presented at scientific conferences.

## ACKNOWLEDGEMENT

The authors would like to thank Angélique Roy for their assistance in developing the literature search strategy.

## STATEMENT OF ETHICS

According to the Health Sciences Research and Ethics Board (HSREB) at Queen’s University, formal ethics approval is not required for this study as no new data will be created or analyzed. The publication of this protocol is part of the research team’s commitment to Open Science and research transparency (39). The publication of this protocol has the potential to help researchers and trainees develop protocols to advance research asking similar questions in different conditions and populations.

## AUTHOR CONTRIBUTIONS

**Ryan H. Kirkpatrick:** conceptualization, methodology, validation, formal analysis, investigation, data curation, writing - original draft, writing - review & editing. **Kristina Nazzicone:** conceptualization, methodology, validation, formal analysis, investigation, data curation, writing - original draft, writing - review & editing. **Aleksandar Biorac:** validation, investigation, data curation, writing - review & editing. **Anneke R. Froentjes:** validation, investigation, data curation, writing - review & editing. **Sonja Molin:** conceptualization, investigation, supervision, writing - review & editing. **Sarah Simpson:** conceptualization, investigation, supervision, writing - review & editing.

## DATA AVAILABILITY

Data sharing is not applicable to this study as no new data will be created or analyzed.

